# Rapid, reliable, and cheap point-of-care bulk testing for SARS-CoV-2 by combining hybridization capture with improved colorimetric LAMP (Cap-iLAMP)

**DOI:** 10.1101/2020.08.04.20168617

**Authors:** Lukas Bokelmann, Olaf Nickel, Tomislav Maricic, Svante Pääbo, Matthias Meyer, Stephan Borte, Stephan Riesenberg

## Abstract

Efforts to contain the spread of SARS-CoV-2 have spurred the need for reliable, rapid, and cost-effective diagnostic methods which can easily be applied to large numbers of people. However, current standard protocols for the detection of viral nucleic acids while sensitive, require a high level of automation, sophisticated laboratory equipment and trained personnel to achieve throughputs that allow whole communities to be tested on a regular basis. Here we present Cap-iLAMP (capture and improved loop-mediated isothermal amplification). This method combines a hybridization capture-based RNA extraction of non-invasive gargle lavage samples to concentrate samples and remove inhibitors with an improved colorimetric RT-LAMP assay and smartphone-based color scoring. Cap-iLAMP is compatible with point-of-care testing and enables the detection of SARS-CoV-2 positive samples in less than one hour. In contrast to direct addition of the sample to improved LAMP (iLAMP), Cap-iLAMP does not result in false positives and single infected samples can be detected in a pool among 25 uninfected samples, thus reducing the technical cost per test to ~1 Euro per individual.

## Introduction

The recent global outbreak of coronavirus disease 2019 (COVID-19) has led governments to take drastic measures to contain the spread of the severe acute respiratory syndrome coronavirus 2 (SARS-CoV-2). Most affected countries resorted to more or less stringent lockdown measures including restrictions on travel, public gatherings and the closing of institutions such as schools, kindergartens and universities. These restrictions and closings have usually been implemented broadly, affecting the lives of infected and uninfected people alike, as no reliable and economical mass testing approach for decentralized point-of-care identification of infected individuals exists.

Reverse transcription followed by quantitative PCR (RT-qPCR) is the most widely used method to detect RNA viruses such as SARS-CoV-2. However, its need for trained personnel and expensive instrumentation and global shortages of resources for RNA purification has spurred search for viable alternatives. Loop-mediated isothermal amplification (LAMP) can rapidly amplify small target nucleic acid sequences under isothermal conditions (Notomi et al. 2000) and has been applied for molecular diagnostics (Wong et al. 2018). The reaction requires four to six primers and produces concatemers of double-stranded amplification products. These can be detected directly, using intercalating dyes (e.g. SYBR green, SYTO-dyes), its reaction with triphenylmethane dye precursors and acid hydrolysis (Miyamoto et al. 2015; Trinh and Lee 2019), or using cleavage with CRISPR enzymes coupled with lateral flow color detection of the cleavage product (Broughton et al. 2020). However, these methods require opening of the tube after the reaction, thus posing the threat of cross-contaminating future reactions with the amplified product. Amplification can also be detected indirectly by hydroxynaptholblue or phenol red based detection of the release of protons and/or pyrophosphate generated during DNA synthesis (Tanner et al. 2015). Recently, a number of studies explored ways to detect SARS-CoV-2 RNA using RT-LAMP (Lamb et al. 2020; Yang et al. 2020; Zhang et al. 2020) but they required time consuming RNA isolation steps before the reaction. There have also been attempts to add sample directly into the reaction without prior purification (Buck et al. 2020; Dao Thi et al. 2020). However, it was noted that the pH of nasopharyngeal swab samples often varies and can adversely affect readouts.

Here we describe a method to detect SARS-CoV-2 RNA of a single infected individual within a bulk sample comprised of up to 26 individual patient samples by combining a hybridization-capture-based RNA extraction approach with smartphone app-assisted colorimetric detection of RT-LAMP products, a procedure that can be performed in less than one hour (Figure 1A).

**Figure 1:**
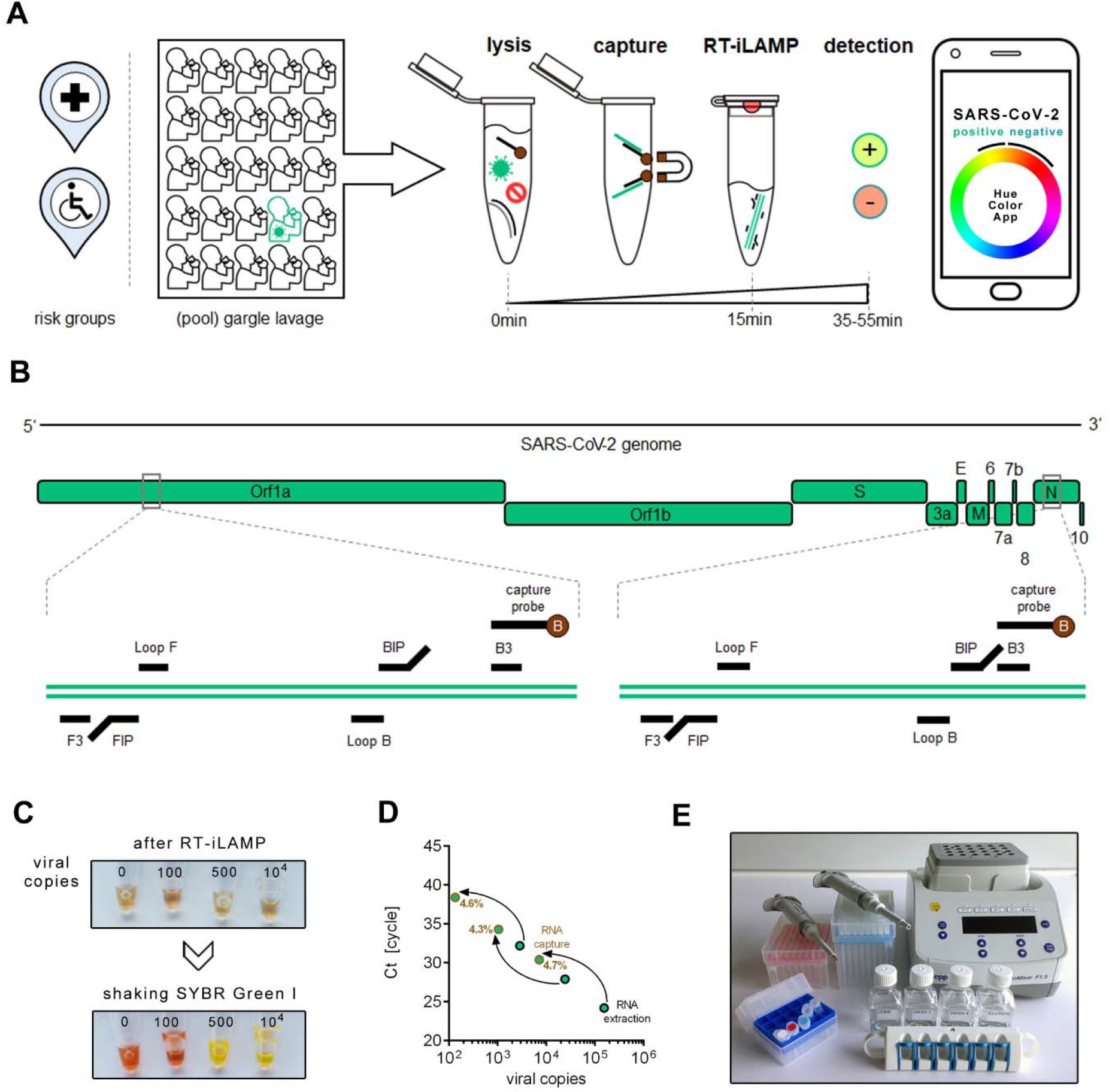
Cap-iLAMP to detect SARS-CoV-2. (A) Workflow of Cap-iLAMP involves collecting and optionally pooling up to 26 gargle lavage samples, followed by combined lysis, target RNA enrichment and improved LAMP (iLAMP). Objective color hue values can be obtained using any freely available ‘camera color picker’ application on a smartphone. (B) Positions of primers and biotinylated capture oligonucleotides targeting the viral Orf1a and N gene on the SARS-CoV-2 genome. (C) Color change induced by dissolving a drop of SYBR green I in the lid of the tube after iLAMP reaction with different input copy numbers of synthetic viral RNA. (D) Capture efficiency was estimated relative to automated silica-based RNA extraction based on copy number estimates for RT-qPCR assay targeting the SARS-CoV-2 E gene. (E) Equipment necessary for Cap-iLAMP: Pipettes and pipette tips, pre-mixed reagents (including iLAMP master mix and capture bead suspension), stable buffers (lysis/binding buffer,wash buffer, low salt buffer, elution buffer), a magnetic rack and a thermoblock.

We compared the sensitivity of the method to standard extraction RT-qPCR protocols in a diagnostic lab and validate its performance on 287 gargle lavage samples from a hospital, a nursing home previously affected by COVID-19, and round robin samples from a reference institution of the German Medical Association.

## Results

### Development of Cap-iLAMP

We evaluated three published RT-LAMP primer combinations targeting either the Orf1a gene or the N gene of the SARS-CoV-2 genome (Lamb et al. 2020; Zhang et al. 2020) using a dilution series of synthetic viral RNA (Twist Biosciences, San Francisco, CA, USA) and chose the two most sensitive primer sets (CV1-6 and CV15-20, Supp. Table 1) to detect the Orf1a gene and the N gene for further testing (Figure 1B, Supp. Figure 1, Supp. Table 1). Using both primer combinations, we detected 500 synthetic viral RNA copies after 25-30 minutes incubation at 65°C as measured by fluorescence real time RT-LAMP (Supp. Figure 1A and C). Combining the primers for the Orf1a gene and the N gene did not increase sensitivity (Supp. Figure 1D).

Amplification in LAMP reactions is often detected colorimetrically by a pH sensitive dye that changes color when extensive DNA synthesis lowers the pH of the reaction (Tanner et al. 2015). As noted in a previous study (Dao Thi et al. 2020), biological samples such as nasopharyngeal swab eluates may change the pH when added to the LAMP reaction directly, leading to false positive results. We found that nasopharyngeal swab eluates tend to be more acidic than gargle lavage samples and that adding gargle lavage directly to a LAMP reaction at a final concentration of 5% leads to false positive results in 4.7% of cases even before the isothermal incubation (Supp. Figure 2A and B).

**Figure 2:**
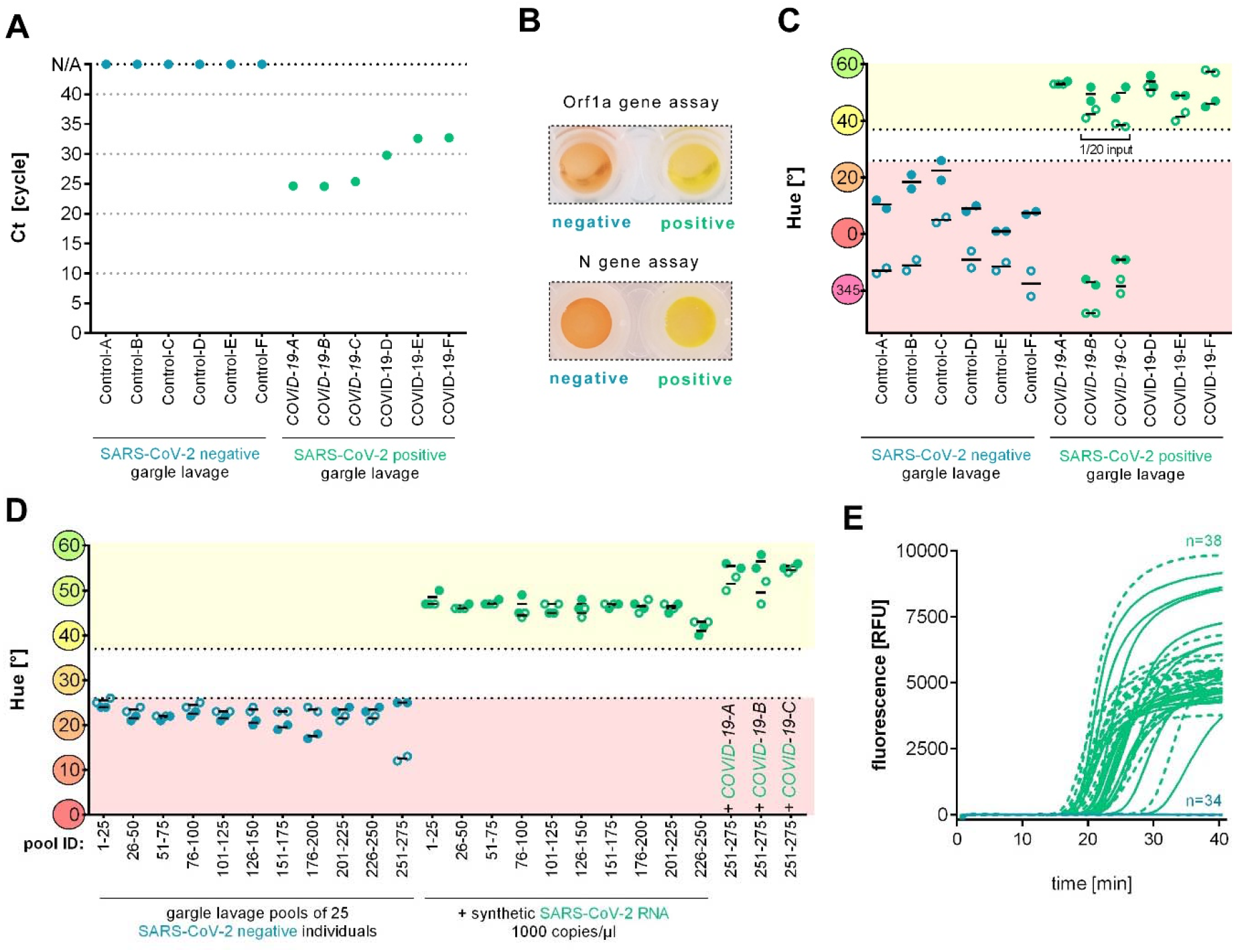
Detection of SARS-CoV-2 in gargle lavage samples. (A) Ct values of individual patient gargle lavage samples obtained via RT-qPCR assay targeting the SARS-CoV-2 E gene. (B) Color of negative and positive control after Cap-iLAMP targeting the Orf1a and N gene of SARS-CoV-2. (C) Hue of individual gargle lavage samples after Cap-iLAMP measured in duplicates. Solid circles indicate values of the Orf1a assay while hollow circles denote values of the N gene assay. (D) Hue of gargle lavage pools after Cap-iLAMP measured in duplicates. Solid circles indicate values of the Orf1a assay while hollow circles denote values of the N gene assay. (E) Amplification plots of LAMP reactions. Dotted lines indicate Orf1a assay while solid lines denote curves of the N gene assay. Data obtained from healthy individuals are shown in blue while SARS-CoV-2 positive patient samples are depicted in green.

It would therefore be preferable to detect the amplification product directly rather than indirectly by a pH change. To achieve this, we deposited a drop of 0.5 microliters of a 10,000x concentrated solution of the dye SYBR Green I to the cap of the tube before the reaction is initiated. Shaking the tube after the isothermal amplification dissolves SYBR Green I, stops the reaction and allows visual detection of SARS-CoV-2 via a color change from orange/red to intense yellow (Figure 1C). The color of the reaction can be objectively quantified as a single numerical hue value, that is insensitive to light intensity changes and can be derived from the red, green, blue (RGB) color model (Cappi et al. 2015; Yin et al. 2019), by using freely available ‘camera color picker’ smartphone apps. We used the ‘Palette Cam’ app (Alexander Mathers, App Store) for extracting RGB values before conversion to hue. An additional advantage is that this detection strategy allows us to include acidifying enhancing enzymes in the LAMP reaction, namely *Tte* UvrD helicase, which prevents unspecific late amplification of artefacts induced by primer interactions (Supp. Figure 3A and B) and thermostable inorganic pyrophosphatase which increases reaction speed (Miyamoto et al. 2015).

However, addition of patient gargle lavage directly to this improved LAMP (iLAMP) reaction promoted false positives (13.5 %) (Supp. Figure 4A). Presumably, this unspecific amplification is due to DNA from the oral microbiome, food or host cells as it can be prevented by prior λ exonuclease treatment that preferentially digests 5’-phosphorylated DNA leaving non-phosphorylated primers and iLAMP product intact (Supp. Figure 4B). Because, gargle lavages from known infected patients did result in false negatives for all seven samples we employed an initial Quick extract (Lucigen, Middleton, WI, USA) lysis (Ladha et al. 2020) to release more viral RNA from the capsid. Even when these optimized reaction conditions were used, reactions resulted in one false negative out of the seven gargle lavages from infected patients and the single SARS-CoV-2 negative gargle lavage sample was false positive (Supp. Figure 4B). We therefore employed a rapid (15min) bead-capture enrichment purification akin to mRNA isolation, using two oligonucleotides flanking the RT-LAMP target sites (Figure 1B, Supp. Table 1) immobilized on paramagnetic beads. This step eliminates non-target nucleic acids and other unwanted components in the biological samples but also concentrates the viral RNA. Comparing the Ct-values of RT-qPCR targeting the E gene (Corman et al. 2020) after silica-based RNA-extraction with hybridization capture targeting the E gene for the same volume of gargle lavage suggests a capture efficiency of roughly 5% (Figure 1D). To allow comparison to RT-qPCR we captured viral RNA with a biotinylated probe for the E gene and not for the Orf1a and N gene as used for RT-iLAMP. When RNA is concentrated from 500μl gargle lavage to 25μl final volume and 10μl input volume is used in iLAMP, this results in a detection limit of 5-25 viral genome copies per μl of sample before capture.

The reagents required for the iLAMP reaction can be pre-mixed and freeze-thawed at least twice. Cap-iLAMP could be performed at point-of-care as no bulky equipment and only pipettes, a thermoblock, and a magnetic rack are needed (Figure 1E).

### Detection of SARS-CoV-2 in patient gargle lavage samples

We used Cap-iLAMP to test 287 gargle lavage samples from hospital patients and elderly nursing home inhabitants and employees either individually or in pools. These samples had previously been tested by a RT-qPCR assay targeting the SARS-CoV-2 E gene (Maricic et al. 2020) and showed either no amplification or Ct values ranging from 24.6 to 32.7 (Figure 2A).

Cap-iLAMP targeting the SARS-CoV-2 Orf1a gene or the N gene results in an orange/red and intense yellow color for SARS-CoV-2 RNA negative and positive samples, respectively (Figure 2B). Of the 12 samples which we tested individually (Figure 2C), six had been previously tested positive in the RT-qPCR assay. Scoring the color of the iLAMP reactions using a smartphone ‘camera color picker’ app shows that hues for SARS-CoV-2 negative and positive samples are clearly separated, below 26° for the former and above 37° for the latter, respectively. All six negative samples were correctly identified as negative in the Cap-iLAMP assays targeting the Orf1a and the N gene. Of the six samples that were SARS-CoV-2 positive in the RT-qPCR assay, four were positive in the Cap-iLAMP assay while the remaining two were false negative. When one twentieth of input volume were used they were positive, suggesting that some residual inhibition originating either from the biological sample or from lysis/binding buffer carryover exists in these extracts.

To investigate whether it is possible to detect single infected individuals in pools of gargle lavage samples, we created eleven pools of 25 patient samples each, all of which had been tested negative in RT-qPCR assay and in the Cap-iLAMP assays for the Orf1a and the N gene (Figure 2D). In order to determine if components of the pooled gargle lavage still inhibit the RT-LAMP reaction after capture, we took subsamples of the 10 negative pools and added 1000 copies/μl of artificial viral RNA before Cap-iLAMP. All 10 pools were positive in both Cap-iLAMP assays, showing that there was no substantial inhibition in these extracts after capture.

To investigate whether it is possible to detect a single infectious individual within a pool, we added different single positive patient samples (Ct < 26) to three pools of healthy individuals so that 1/26 (3.8%) of the final volume was composed of the infected sample. In all three cases, the Cap-iLAMP assays targeting the Orf1a and the N gene were positive, demonstrating that a single infectious individual can be detected in a pool of 26 samples. The two SARS-CoV-2 positive samples previously found to be inhibited when tested individually (Figure 2C) were correctly identified as positive when pooled (Figure 2D), potentially because inhibition is rare and is diluted out in the pool. An additional wash step after hybridization capture should thus be applied for individual sample diagnostic testing.

All tested gargle lavages from single healthy individuals (n=6) and 11 pools of 25 healthy individuals (n=275) correctly tested negative for both the Orf1a gene and N gene in the Cap-iLAMP assay (Figure 2C and E), indicating that false positive results which were sometimes observed when samples are added directly into the iLAMP reaction (Supp. Figure 4A) are effectively prevented by the hybridization capture extraction.

### Detection of SARS-CoV-2 in heat-inactivated cell-culture supernatants

Finally, we validated Cap-iLAMP against a set of seven reference samples of heat-inactivated cell-culture supernatants provided by the “INSTAND” reference institution of the German Medical Association (Duesseldorf, Germany) for round robin testing. These contained various amounts of SARS-CoV-2 (ring 59, 61, 63 and 64), different coronaviruses (ring 60 HCoV OC43 and ring 65 HCoV 229E) or no virus (ring 62) and had previously been tested via RT-qPCR (Maricic et al. 2020) (Figure 3A). To prevent sporadic inhibition as observed for some gargle lavage samples, the first wash step after capture hybridization was performed twice. As shown in Figure 3B, all four SARS-CoV-2-containing samples were correctly and consistently identified by both the Orf1a and N gene assays while all samples devoid of virus or containing a different coronavirus were correctly classified as being negative.

**Figure 3:**
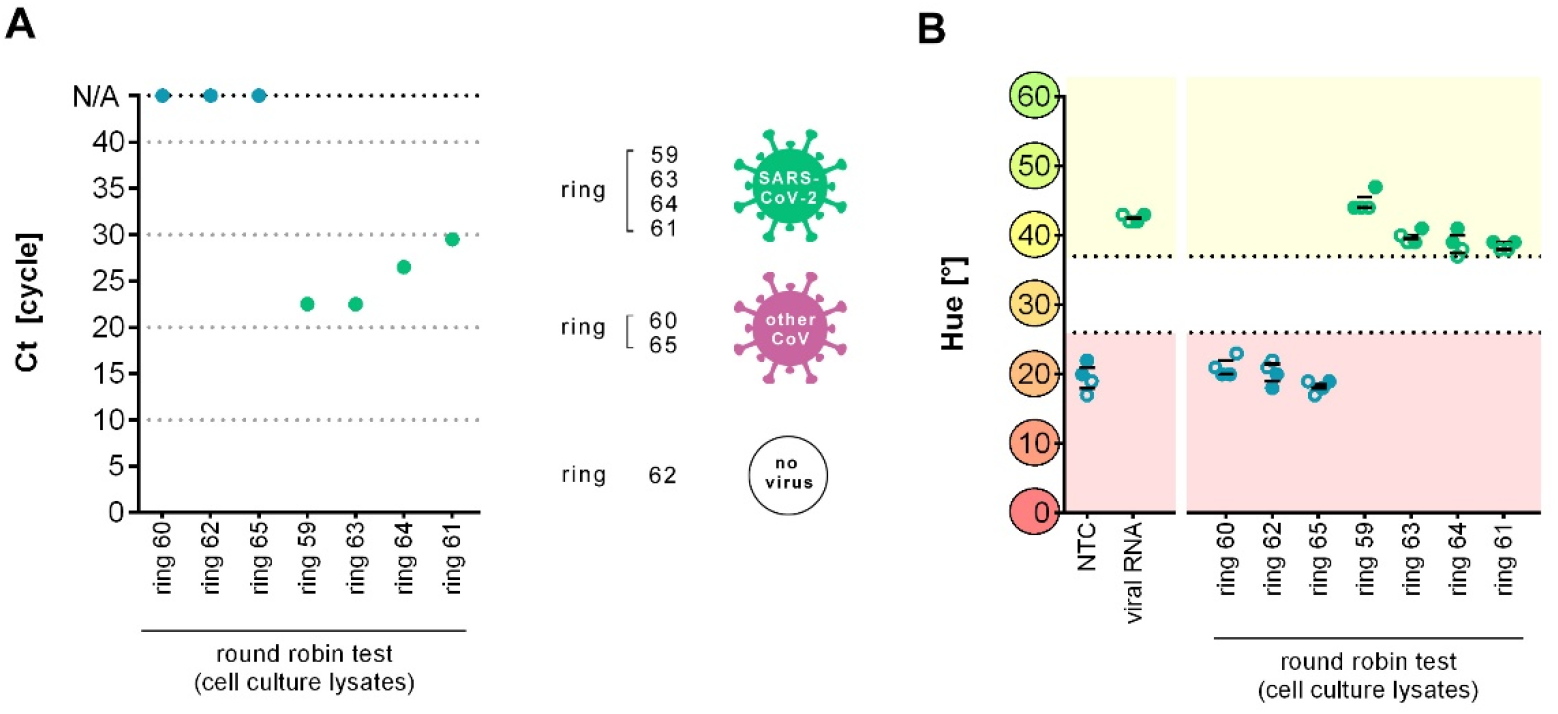
Detection of SARS-CoV-2 in cell culture supernatants provided for assay validation. (A) Ct values of individual cell culture supernatant samples obtained via RT-qPCR assay targeting the SARS-CoV-2 E gene. (B) Hue of individual cell culture supernatant samples after Cap-iLAMP measured in duplicates. Solid circles indicate values of the Orf1a assay while hollow circles denote values of the N gene assay. Data obtained from healthy individuals are shown in blue while SARS-CoV-2 positive patient samples are depicted in green.

## Discussion

We have established Cap-iLAMP as a reliable and rapid diagnostic test for SARS-CoV-2. Our method overcomes problems associated with standard RT-LAMP pH-dependent colorimetric detection which is prone to false positives due to pH-variability in gargle lavages and off-target amplification. Gargle lavage cause no discomfort and is thus more suitable than pharyngeal and nasal swabs for frequent and repeated testing of apparently healthy individuals (Saito et al. 2020). They have the additional advantage that they do not expose the person collecting samples to any risk of infection.

In contrast to RT-qPCR approaches, no complicated equipment such as qPCR machines is required. Removal of inhibitors and enrichment of target nucleic acids by on-bead capture, combined with a double assay of two SARS-CoV-2-specific target sites and control reactions to detect inhibitor carryover and contaminated reagents ensure accuracy. As quality controls we recommend dedicated water negative controls and sample-specific inhibition/positive controls containing sample and synthetic viral RNA for each the Orf1a and the N gene assay. A scheme for evaluating the results is shown in Supp. Table 2.

Due to the removal of non-target nucleic acids and the inclusion of LAMP enhancing enzymes in Cap-iLAMP, we did not detect false positive results, which were often reported in previous LAMP-based detection methods (Teng et al. 2007; Abbasi et al. 2016; Hardinge and Murray 2019). The inclusion of dye in the cap of the tube allows to keep the tube closed for detection after amplification thus preventing the cross-contamination of future assays. We quantified color as a single numeric hue value using a freely available smartphone app, which allows objective point-of-care detection without the ambiguity of an individual’s perception of color. Yin et al. (Yin et al. 2019) already described application of hue for hydroxynaptholblue color scoring that is in need of a custom-made ‘smart cup’ reaction tube holder as well as an app that is not freely available. Consequently, Cap-iLAMP outperforms other published SARS-CoV-2 detection methods (Broughton et al. 2020; Buck et al. 2020; Corman et al. 2020; Dao Thi et al. 2020) in some key aspects as shown in Table 1.

**Table 1:** Comparison of SARS-CoV-2 detection methods. Ten important aspects of different published SARS-CoV-2 detection methods and Cap-iLAMP are compared and ranked optimal (green), sub-optimal (light green), and critical (orange).

|  | RNA extraction / RT-qPCR | DETECTR RT-LAMP / Cas12a | Direct sample to RT-LAMP | Cap-RT-iLAMP |
| --- | --- | --- | --- | --- |
| Reference | Corman et al. | Broughton et al. | Dao Thi et al., Buck et al. | this study |
| Sample-to-result time | 4h | 45min | 30min | 55min |
| Reagent price | \$\$\$ | \$\$ | \$ | \$ |
| Bulky equipment | yes | no | no | no |
| Limit of detection | 1 copy | 10 copies | 100-500 copies | 100-500 copies (eq. to 5-25 copies per μl sample liquid before capture) |
| Inhibitor removal | yes | no | no | yes |
| Sporadic false positives | no | no | yes | no |
| Open-lid cross-contamination | no | yes | no | no |
| Assay result detection | fluorescence (FAM) | colorimetric (FAM) | colorimetric (pH-dye) | colorimetric (cap SYBR Green I) |
| Helicase or PPase addable | not applicable | yes | no | yes |
| Assay result scoring | quantitative Ct value | lateral flow | subjective visual scoring | smartphone → quantitative hue |

The Cap-iLAMP assay passed an official round robin test used to evaluate testing accuracy across diagnostic laboratories. We show that it is possible to detect a single infected individual in a pool of 26 samples, opening the prospect of rapid screening of large numbers of people in public decentralized settings such as in schools, hospitals, airports, prisons, retirement homes or border crossings. This also allows daily testing of people at risk or working in critical infrastructure.

The Cap-iLAMP method can be readily applied to numerous human respiratory pathogens, plant pathogens, animal pathogens, food-borne diseases, viruses, protozoan parasites, and fungi, for which primer combinations have been developed (Wong et al. 2018). These include the eight diseases prioritized by the WHO (as of July 2020) that pose the greatest public health risk due to their epidemic potential and/or whether there is no or insufficient countermeasures (Kurosaki et al. 2010; Fukuma et al. 2011; Osman et al. 2013; Oloniniyi et al. 2017; Huang et al. 2018; Ma et al. 2019). For testing in remote areas or developing countries master mixes for iLAMP could potentially be lyophilized for transportation or long term storage, as shown previously (Chen et al. 2016; Carter et al. 2017).

## Materials and Methods

### Sampling of gargle lavages

Sampling of gargle lavages was done as described previously by our group (Maricic et al. 2020). All individuals included in this study were asked for their voluntary assistance to participate and each individual gave written informed consent before entry into the study. The study was approved by the Ethics Committee of the Saxonian medical chamber (EK-allg-37/10-1). All procedures utilized in this study are in agreement with the 1975 Declaration of Helsinki. 10 ml of sterile water was filled into sterile urine cups. The full water volume was gargled for 10 seconds before spitting it back into the cup. Analyzed anonymized gargle lavages from hospitalized COVID-19 patients and round robin samples (INSTAND) were kept frozen below -80°C for 1-2 months.

### In solution capture purification of SARS-CoV-2 nucleic acids

Per reaction 20μl Dynabeads MyOne Streptavidin C1 bead suspension (Thermo Fisher Scientific, #650.02) are pelleted using a magnetic rack and washed twice with 500μl combined 1x lysis/binding buffer (LysBB: 100 mM Tris-HCl, pH 7.5, 500 mM LiCl, 0.5% LiDS, 1 mM EDTA, 5 mM DTT). Washed beads are then resuspended in 100μl 1x LysBB before addition of 20pmol CV2_btn and 20pmol CV16_btn (see Supp. Table 1) (or 10pmol E_Sarbeco_R2_btn for the RT-qPCR assay) and rotation for 10min at room temperature. Beads are pelleted and supernatant is discarded before resuspension in 200μl alkaline wash solution (125mM NaOH, 0.1% (v/v) Tween-20) and 5min incubation at room temperature. Beads are pelleted with a magnetic rack and supernatant is discarded. Beads are then washed once with 200μl 1x LysBB before resuspension in 500μl 2x LysBB. Prepared bead suspension can be stored at 5°C or used directly.

To 500μl ready-made bead suspension an equal volume of patient gargle lavage (sample), water (negative control) or sample with 500k copies of artificial SARS-CoV-2 RNA (Twist Biosciences, #102024) (positive and inhibition control) is added and mixed by pipetting. The reaction is incubated at 55°C for 10min and then placed on a magnetic rack. The supernatant is discarded and the beads are thoroughly resuspended in 200μl wash buffer (20 mM Tris-HCl, pH 7.5, 500 mM LiCl, 1 mM EDTA). Beads are pelleted and supernatant is discarded, repeat this step once for individual or inhibited samples. Wash beads once with 200μl low salt buffer (20 mM Tris-HCl, pH 7.5, 200 mM LiCl, 1 mM EDTA) and discard supernatant. Beads are resuspended in 25μl elution buffer (20 mM Tris-HCl, pH 7.5, 1 mM EDTA) and RNA is eluted by heating to 60°C for 2min. Transfer the supernatant to a fresh 0.2ml strip tube or use directly as input for RT-iLAMP or RT-qPCR.

### RT-iLAMP

For each sample two assays targeting either the Orf1a or N gene are performed. Primer mixes for both assays are prepared in bulk for quick reaction assembly (Supp Table 3). 20μl of RT-iLAMP master mix containing either the Orf1a (CV1-6) or N gene (CV15-20) primer sets (Supp. Tables 4) and 10μl of capture eluate are combinded to obtain a final reaction volume of 30μl (1x WarmStart® Colorimetric LAMP Master Mix (NEB, Ipswich, MA, USA, #M1800L), 1mM ATP, 1μM SYTO 9, 1x primer mix, 0.1ng/|μl Tte UvrD Helicase (NEB, #M1202S), 0.05U/|μl thermostable inorganic pyrophosphatase (NEB, #M0296L), 0.4U/μl Protector RNase inhibitor (Sigma Aldrich, St. Louis, MI, USA, # 03335402001)). For color reactions, 0.5μl SYBR green I is applied to the lid of the tube without getting into contact with the reaction liquid. For transport or preparation of large numbers of reactions it is also possible to immobilize the dye by drying for 15min at 70°C in an oven. If amplification curves should be recorded on a qPCR machine, no SYBR green I is needed. The reaction is incubated at 65°C for 40min. If SYBR green I was applied to the cap of the tube, the reaction is then stopped by shaking and can be evaluated by eye or with any ‘camera color-picker’ app on a smartphone within a minute. We used the ‘Palette Cam’ app to score RGB values and extracted the hue. This can be easily done using a web tool (e.g. https://www.rapidtables.com/convert/color/rgb-to-hsv.html). Hue can also be scored directly when using e.g. ‘Color Grab (color detection)’ app for android or ‘Aurora’ app for iOS.

## Data Availability

Data is available upon request

## Acknowledgements

We thank the management of Diakonische Dienste Leipzig, the personnel and residents of Altenpflegeheim Emmaus in Leipzig, and the personnel of the Department of Laboratory Medicine at Hospital St Georg Leipzig for their cooperation. Funding was provided by the Max Planck Society, the NOMIS foundation, and the ImmunoDeficiencyCenter Leipzig.

## Supplementary Data

**Supplementary Table 1:**
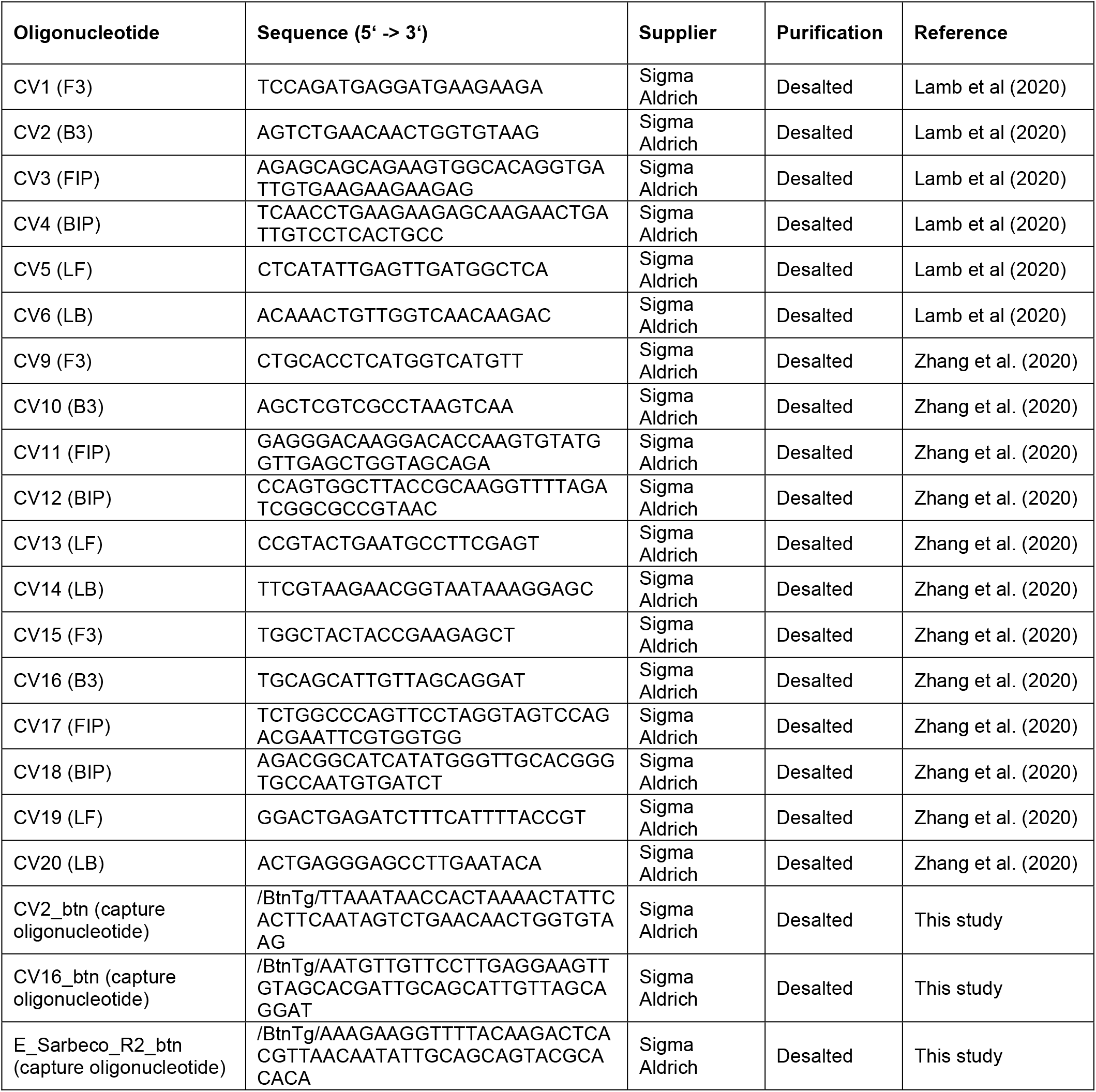
Sequences of oligonucleotides used in this study.

**Supplementary Table 2:** Evaluation of Cap-iLAMP results. For each of the two assays (targeting either the SARS-CoV-2 Orf1a or N gene), a water negative control as well as a sample-specific positive/inhibition control comprised of sample and artificial viral RNA should be included.

| Assay | Patient sample | + Inhibition control | Negative control | RESULT |
| --- | --- | --- | --- | --- |
| Orf1a | + | + | - | SARS-CoV-2-positive |
| N | + | + | - |  |
| Orf1a | - | + | - | Presumptive positive |
| N | + | + | - |  |
| Orf1a | + | + | - |  |
| N | - | + | - |  |
| Orf1a | - | + | - | SARS-CoV-2-negative |
| N | - | + | - |  |
| Orf1a | + | + | + | QC failure |
| N | + | + | + |  |
| Orf1a | - | - | - |  |
| N | - | - | - |  |

**Supplementary Table 3:** Composition of a 10x LAMP primer master mix.

| Component | V/μl | Final concentration |
| --- | --- | --- |
| F3 primer (100μM) | 8 | 2μM |
| B3 primer (100μM) | 8 | 2μM |
| FIP primer (100μM) | 64 | 16μM |
| BIP primer (100μM) | 64 | 16μM |
| LF primer (100μM) | 16 | 4μM |
| LB primer (100μM) | 16 | 4μM |
| Water | 224 | - |
| **Total** | **400** |  |

**Supplementary Table 4:** Composition of the iLAMP master mix for a 30μl reaction.

| Component | V/μl | Concentration in 30μl reaction |
| --- | --- | --- |
| WarmStart® Colorimetric LAMP Master Mix (2x) (NEB) | 15 | 1x |
| Primer mix (10x) | 3 | 1x |
| ATP (100mM) | 0.3 | 1mM |
| SYTO-9 (100μM) | 0.3 | 1μM |
| Tte UvrD helicase (20ng/μl) | 0.15 | 0.1ng/μl |
| Protector RNase inhibitor (40U/μl) (Sigma Aldrich) | 0.3 | 0.4U/μl |
| Thermostable inorganic pyrophosphatase (2U/μl) (NEB) | 0.75 | 0.05U/μl |
| Nuclease free water | 0.2 | - |
| **Total** | **20** |  |

**Supplementary Fig. 1.**
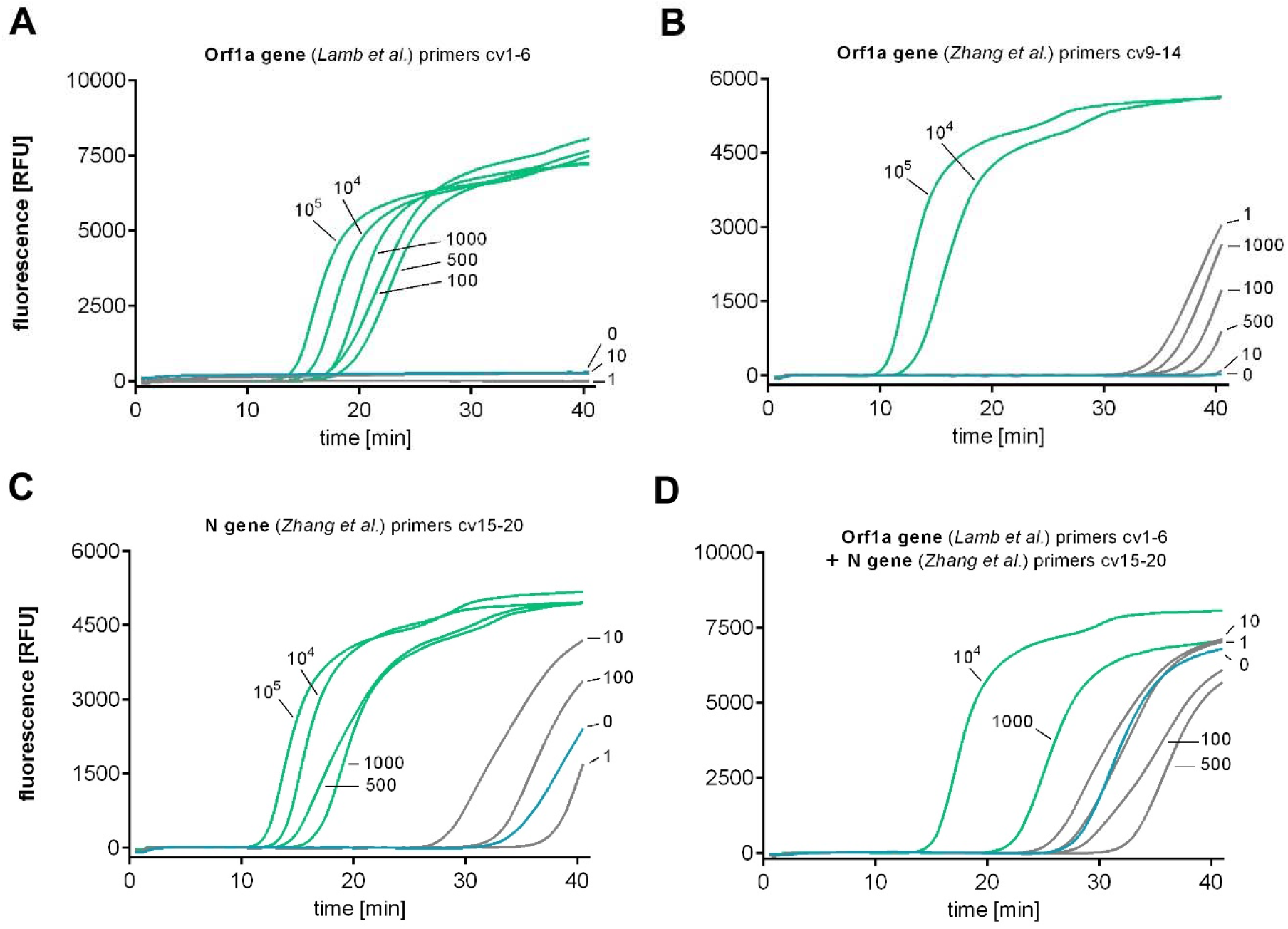
Sensitivity of different primer sets targeting SARS-CoV-2 RNA. (A) Amplification curves of LAMP reactions using primers CV1-6 and various copy numbers of artificial SARS-CoV-2 RNA. (B) Amplification curves of LAMP reactions using primers CV9-14 and various copy numbers of artificial SARS-CoV-2 RNA. (C) Amplification curves of LAMP reactions using primers CV15-20 and various copy numbers of artificial SARS-CoV-2 RNA. (D) Amplification curves of LAMP reactions using two primer sets (CV1-6 + CV15-20) and various copy numbers of artificial SARS-CoV-2 RNA. Amplification proceeded at 65°C.

**Supplementary Fig. 2.**
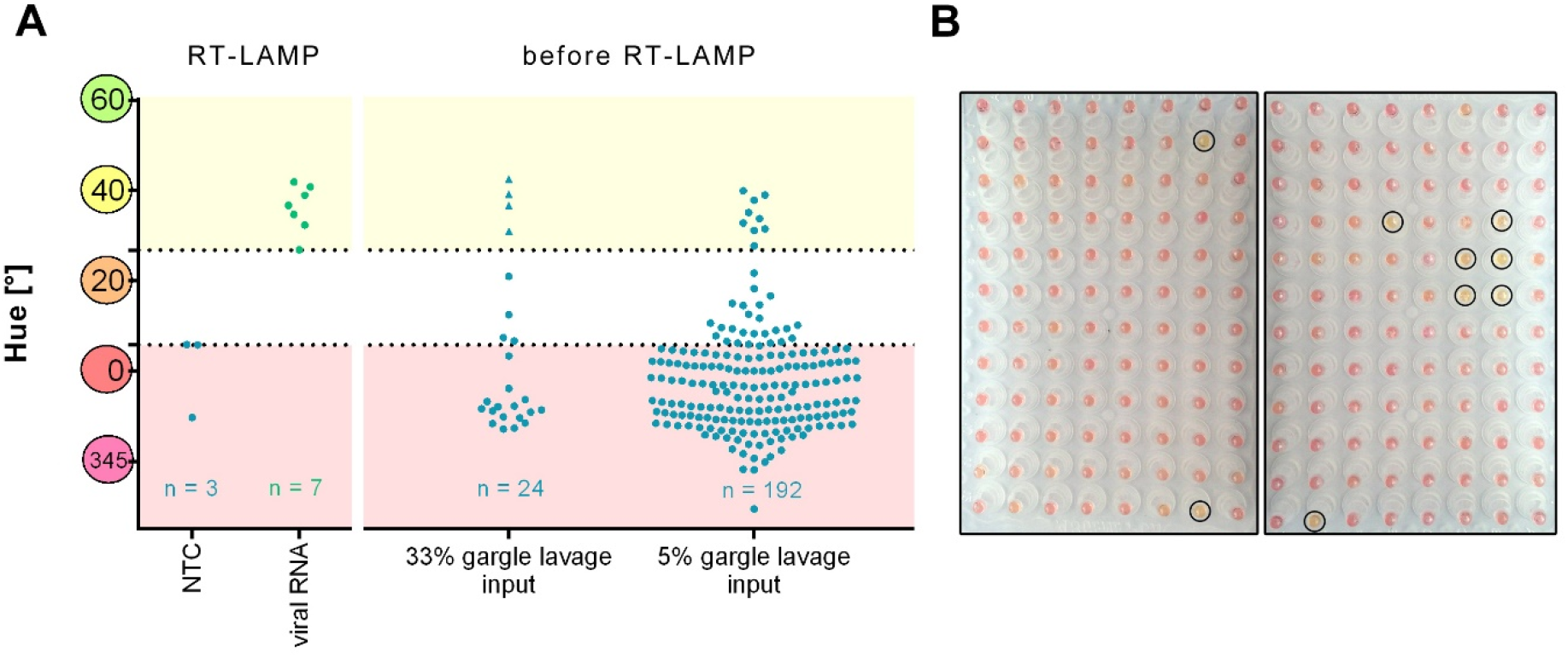
pH variability in diagnostic samples can lead to false positives. (A) Hues measured for negative and positive control reactions after RT-LAMP (left) and patient gargle lavage (GL) samples added directly to the LAMP reaction in different ratios (33% and 5%) before performing RT-LAMP. Triangles mark nasopharyngeal swab eluate samples. (B) LAMP reaction mixes including directly added gargle lavage samples before RT-LAMP. Samples passing the positive hue threshold are marked by circles.

**Supplementary Fig. 3.**
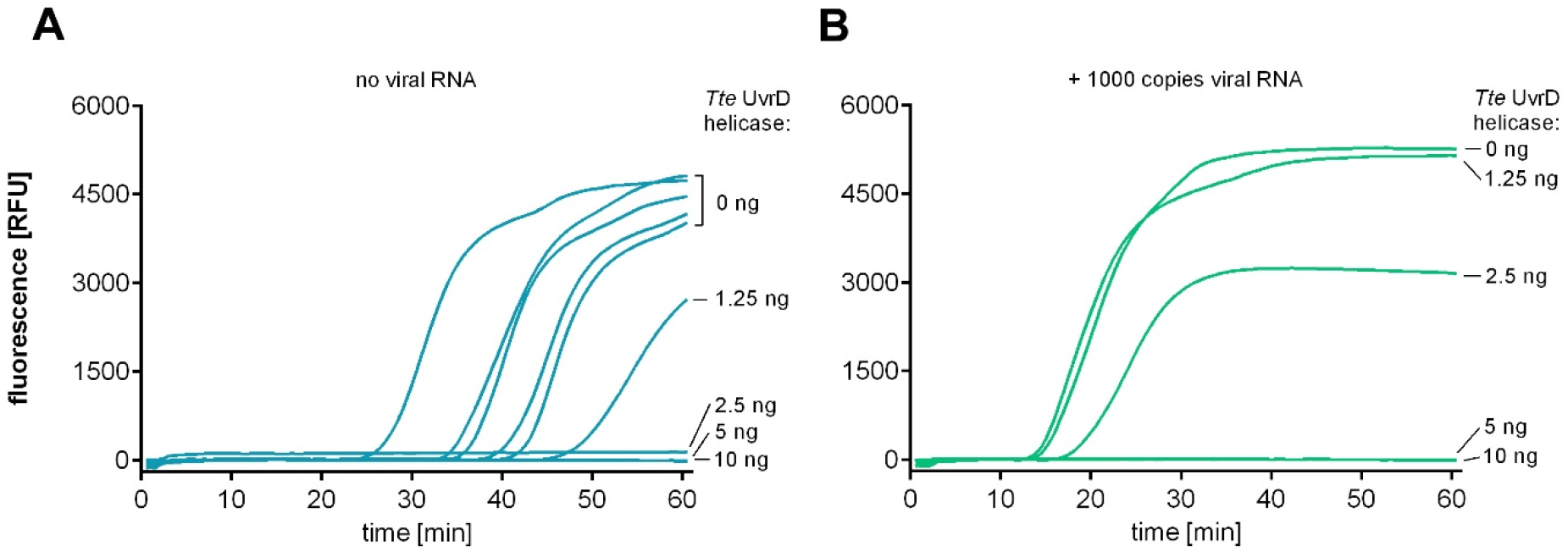
Evaluation of different amounts of *Tte* UvrD helicase in LAMP reactions. Amplification plots of negative controls (left panel) and positive controls containing 1000 copies of artificial SARS-CoV-2 RNA (right panel). Amount of *Tte* UvrD helicase in 20μl reaction are indicated on the right. The primer set used targeted the N gene (CV15-20), the assay proceeded at 65°C.

**Supplementary Fig. 4.**
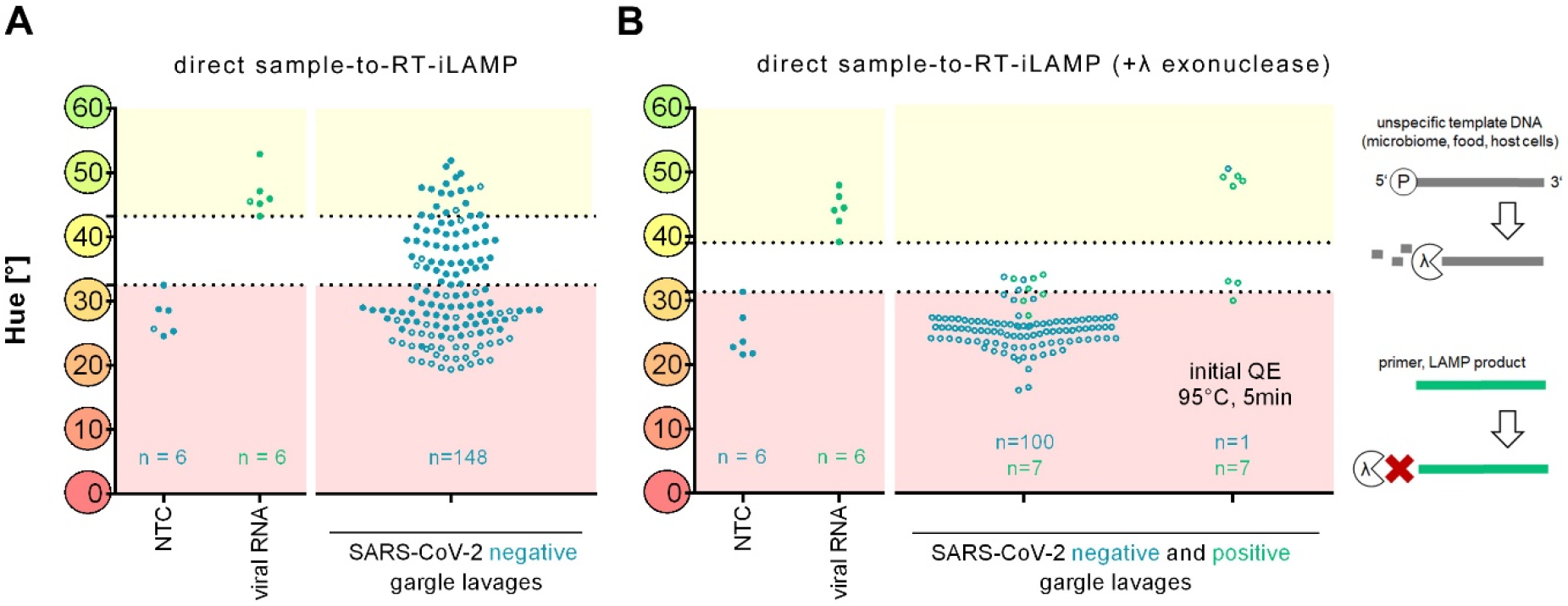
Direct LAMP of patient gargle lavage samples can lead to false positives and false negatives. (A) Hues measured for negative (n=6) and positive (n=6) controls after RT-iLAMP reactions (left side) and after RT-iLAMP of SARS-CoV-2-negative (n=148) gargle lavage samples directly added to the reaction mix. (B) Hues measured after RT-iLAMP reactions preceeded by a λ exonuclease (NEB) digestion step (5U per reaction for 10-15min at 25°C) for negative (n=6) and positive (n=6) controls (left side), SARS-CoV-2-negative (n=100) and SARS-CoV-2-positive (n=7) gargle lavage samples directly added to the reaction mix (middle) and SARS-CoV-2-negative (n=1) and SARS-CoV-2-positive (n=7) gargle lavage samples directly added to the reaction mix digested with QuickExtract (QE) and heat inactivated for 5min at 95°C before LAMP (right). Schematic drawing of λ exonuclease digestion shown on the right. Primers and LAMP reaction products do not possess a 5’-phosphate and are thus not a target for λ exonuclease digestion. All reactions contained 10μg T4 gene 32 protein (NEB) as it could alleviate inhibition of some gargle lavages in initial experiments.

